# An LC-immuno-MRM-MS Winterplex Method Development Framework for Respiratory Viral Screening

**DOI:** 10.1101/2025.07.18.25331758

**Authors:** Bart Van Puyvelde, François E. Dufrasne, Sarah Denayer, Anna Parys, Tanika Van Mulders, Marijn Van Hulle, Dieter Deforce, Cyril Barbezange, Koen De Cremer, Steven Van Gucht, Johannes PC Vissers, Maarten Dhaenens

**Author notes:** Corresponding author: Maarten Dhaenens –.

## Abstract

Reliable large-scale testing for respiratory viruses, including influenza viruses, coronaviruses, such as SARS-CoV-2, and respiratory syncytial virus (RSV) is essential for both endemic surveillance and pandemic response. While RT-qPCR remains the current gold standard, the COVID-19 pandemic highlighted the need for alternative, scalable detection platforms. Although alternative approaches like liquid chromatography coupled with mass spectrometry (LC-MS) are well-suited for viral multiplexing, they have yet to undergo clinical validation. Here, we present a quantitative immuno-multiple reaction monitoring (iMRM) assay, called Winterplex, for the simultaneous detection of influenza A, influenza B, RSV, and SARS-CoV-2, each targeted via two proteotypic peptides per virus. The method was validated according to ISO standards for *in vitro* diagnostics and benchmarked against RT-qPCR. To enhance future pandemic preparedness, urgent investment is needed to translate and expand multiplexed MS-based diagnostic methodologies, which offer the flexibility to incorporate a wide range of targets, making them ideally suited for rapid and adaptable responses to emerging viral threats.

## INTRODUCTION

Seasonal respiratory infections, particularly during winter months, pose a significant challenge to public health and society. In the aftermath of the Covid-19 pandemic, influenza and respiratory syncytial virus (RSV) have resurged, highlighting the evolving dynamics of viral transmission ^1^. The temporary suppression of these pathogens due to widespread non-pharmaceutical interventions created a gap in immunity, leading to more severe and widespread outbreaks as restrictions were lifted ^2^. These resurgences place considerable pressure on healthcare systems by increasing hospitalizations and straining medical resources, while also disrupting economic activity. Rapid and scalable diagnostic testing is essential for early detection and containment, enabling timely interventions such as personal protective equipment, targeted patient isolation, optimized vaccine deployment, and the use of antiviral therapies. Strengthening diagnostic and surveillance infrastructure is critical to mitigate the recurrent impact of seasonal respiratory viruses ^3^. The need for effective surveillance has become even more pressing with the emergence of novel viral strains, notably the highly pathogenic avian influenza (HPAI) clade 2.3.4.4.b., which poses a heightened risk of zoonotic transmission and raises concerns about novel outbreaks ^4–6^. In the United States, a recent surge of HPAI among poultry flocks and cattle has already contributed to egg shortage and highlights the importance of influenza A subtype testing for clinical specimens, to detect infections early and implement effective interventions ^7,8^. Addressing these challenges requires the development of sensitive, specific, flexible and comprehensive diagnostic tools for the detection of respiratory viruses across diverse clinical and environmental settings.

Currently, real-time quantitative polymerase chain reaction (RT-qPCR) remains the gold standard for viral diagnostics due to its high sensitivity, even at low viral loads ^9^. During the SARS-CoV-2 pandemic, global shortages of RT-qPCR reagents highlighted the urgent need for alternative diagnostic methodologies to ensure system resilience ^10^. These shortages were primarily driven by (i) disruptions in commercial supply chains due to lockdowns and (ii) the unprecedented testing demands imposed by health authorities for effective monitoring and control. In response, orthogonal approaches such as liquid chromatography– mass spectrometry (LC-MS) were developed, offering increased sample throughput and reliable detection using reagents distinct from those required for RT-qPCR. Early work by several independent groups demonstrated that LC-MS can complement existing diagnostic technologies, offering a robust and scalable alternative that enhances viral surveillance and ensures preparedness for future outbreaks ^11–16^.

To achieve viral detection sensitivity comparable to RT-qPCR, immuno-enrichment strategies targeting specific viral peptides have been employed. More specifically, the use of anti-peptide antibodies to selectively enrich target peptides from complex biological samples, such as nasopharyngeal (NP) swabs and saliva, has been a key advancement. This method, known as immuno-Multiple Reaction Monitoring (iMRM), combines targeted peptide enrichment with LC-MRM analysis to ensure high specificity and accurate quantification of viral load in many different matrices ^17–20^. Unlike RT-qPCR, which relies on amplifying viral genetic material, iMRM directly measures viral proteins, eliminating the risk of amplification bias, primer-dimer artifacts, and other technical errors. In turn, this intrinsically makes peptide detection much more quantitatively accurate. Additionally, while RT-qPCR is highly sensitive, it can detect residual viral RNA for certain viruses long after the active infection has been resolved, leading to potential false-positive results ^21,22^. In contrast, protein-based detection by LC-MS is considered to provide a more accurate measurement of active infection, reducing the likelihood of misclassification due to lingering genetic material.

Another advantage of LC-MS is its ability to handle pooled samples without significant sensitivity loss when paired with peptide immuno-enrichment. This is particularly beneficial in large-scale screenings, such as wastewater surveillance or monitoring incoming transcontinental flights for emerging viral threats^23^. While pooling in RT-qPCR dilutes the genetic material thereby proportionally compromising detection, iMRM remains highly sensitive, optimizing resource efficiency and reducing diagnostic costs in low-prevalence scenarios ^19^.

Beyond infectious disease detection, iMRM has broad applications in monitoring clinically relevant biomarkers across various disease areas. It has been employed for the quantification in plasma and serum of acute inflammation markers such as C-reactive protein (CRP) and serum amyloid A (SAA), cardiovascular risk markers including apolipoproteins ApoA-I and ApoB, and even cancer biomarkers such as thyroglobulin for thyroid cancer detection ^24–28^. This targeted approach enables precise, reproducible, and multiplexed quantification, making it a powerful tool for disease monitoring, risk assessment, and personalized medicine. When combined with Dried Blood Spots (DBS), iMRM offers a convenient, minimally invasive sampling method suitable for large-scale epidemiological studies, allowing individuals to collect samples at home for longitudinal disease tracking.

In this study, we describe a multiplexed viral respiratory iMRM method, which we termed Winterplex, designed to precisely quantify SARS-CoV-2, influenza A, influenza B, and RSV viral loads from clinical NP swabs. To achieve high specificity, polyclonal anti-peptide antibodies were developed targeting two distinct peptides per virus. Absolute quantification is ensured by combining all target peptides into a concatenated isotope-labeled protein standard, specifically engineered for this project using QconCAT technology. The analytical performance of the method, including selectivity, linearity, carryover, matrix effects, stability, and imprecision was rigorously validated according to the Clinical and Laboratory Standards Institute (CLSI) C64 guidelines, while sensitivity was benchmarked against RT-qPCR cycle threshold (Ct) values ^29^. While the Winterplex method enables detection of distinct viruses, it currently lacks the capability to differentiate between viral subtypes. Therefore, we propose peptide targets that could enable future subtyping, specifically for IAV H1N1 vs H3N2 and an avian influenza strain. By integrating targeted MS with immuno-enrichment, this method provides a scalable and high-throughput diagnostic platform with the potential to enhance respiratory virus surveillance and strengthen pandemic preparedness. A standardized SOP ensures seamless deployment, preventing LC-MS instruments from remaining idle when the next pandemic hits.

## MATERIAL AND METHODS

### Reagents and materials

A ^15^N labeled Cov2MS Quantification conCATamer (QconCAT) synthetic protein comprising several peptide targets of SARS-CoV-2, influenza A/B and RSV (Supplementary Figure 1), was obtained from Polyquant (Cov²MS, Bad Abbach, Germany). Unlabeled (light) and stable isotope labeled (SIL or heavy) peptides for the different peptide targets were obtained from Biosynth (Staad, Switzerland). Upon receipt, the lyophilized peptides were solubilized in 30% acetonitrile (ACN)/0.1% formic acid (FA) and stored at −80 °C. Polyclonal (VLS, SMV, DQL, VII, GVF and MVL) and Monoclonal (AYN and ADE) anti-peptide antibodies coupled to magnetic Dynabeads Protein G (Thermo Fischer Scientific, MA, USA) were sourced from SISCAPA Assay Technologies (Victoria, Canada) and stored at 4–8 °C. Ammonium bicarbonate (ABC) and Tosyllysine Chloromethyl Ketone (TLCK) and 3-[(3-Cholamidopropyl)dimethylammonio]-1-propanesulfonate (CHAPS) were sourced from Sigma-Aldrich (St. Louis, USA). Trypsin/Lys-C mix was sourced from Promega (Madison, Wisconsin, USA).

### Clinical samples

Residual nasopharyngeal samples screened for SARS-CoV-2, influenza B, influenza A and RSV by RT-qPCR were obtained from the Central Biobank Platform Sciensano (Ixelles, Belgium), with approval of the University Hospital Ghent ethics committee (ONZ-2023-0500). The research conducted in this study adheres rigorously to all pertinent regulations, including the criteria outlined by the Declaration of Helsinki, concerning the involvement of human study participants.

### Avian flu sample

A H5N1 strain of 2023, A/Gallus_gallus/Belgium/00548_0001/2023 (H5N1), provided by the Belgian National Reference Laboratory for Avian Influenza (Sciensano), was used in this study. The viral isolate was initially amplified from a brain swab (ECE2) using embryonated chicken eggs. Following amplification, the highly concentrated viral stock was inactivated by treatment with β-propiolactone (BPL). The BPL inactivation was subsequently followed by a dialysis step to remove residual BPL. To confirm complete viral inactivation, the treated viral stock was inoculated into embryonated chicken eggs and passaged; absence of viral replication confirmed successful inactivation. The viral culture, with a quantified HA-titer of 430.5, was analyzed using two approaches: 180 µL of sample was used for untargeted (ZenoSWATH) analysis, while a dilution series of 2.5, 5, 10, 20, 40, 80, and 160 µL was prepared for targeted (MRM) analysis.

### Protein extraction and digestion

180 µL nasopharyngeal swab sample was precipitated by adding 1260 µL (9 volumes) of ice-cold acetone (−20°C), as described previously for SARS-CoV-2 detection ^19^. After centrifugation at 16,000 x g for 10 min at 0°C, the supernatant was discarded and 1 µg of Trypsin/Lys-C mix (Promega) and 91.3 fmol of the QconCAT standard in 100 µL 50 mM ABC buffer was added. This was followed by an incubation step of 30 min at 37°C, to facilitate trypsin digestion. Next, the trypsin digestion was quenched by adding 10 µL of a 0.22 mg/mL Tosyl-L-lysyl-chloromethane hydrochloride in 10 mM HCl solution, followed by a 5-min incubation step at room temperature.

### Peptide enrichment

First, the required amount of Dynabeads Protein G (30 mg/mL), namely 5 µL of 30 mg/mL beads per 1 µg antibody, were washed three times in a 2 mL protein LoBind® Eppendorf tube using a wash buffer composed of 0.03% CHAPS in 1x PBS. After the required amount of polyclonal antibody was added to the bead mixture, the solution was diluted with wash buffer to an antibody concentration of approximately 0.01 mg/mL, followed by end-over-end mixing at 4°C. Following overnight incubation, the supernatant was removed and replaced with 15 mL crosslinking solution (20 mM DMP in 200 mM triethanolamine, pH 8.5), followed by end-over-end mixing at room temperature for 30 min. Again, the tube was placed on a magnet to draw the beads to the side, allowing complete aspiration of the supernatant. To quench the reaction, 15 mL of a 150 mM monoethanolamine solution (pH 9) was added, followed by a 30 min end-over-end incubation at room temperature. The supernatant was then removed while keeping the tube on the magnet. Any residual unbound material was washed away with 15 mL of an acidic wash solution (0.03% CHAPS in 0.1% formic acid). As this step could lead to antibody uncoupling, the tube was immediately removed from the magnet once the beads were fully resuspended through vortexing. To eliminate any remaining acidic residue, the beads were washed twice with wash buffer before being resuspended in 0.03% CHAPS and 0.01% NaN₃ in 1× PBS to a final concentration of 0.1 µg/µL (0.5 µg/µL for DQLLSSSK). The crosslinked antibodies were then stored at 4°C.

Prior to the addition of the antibody-coupled magnetic bead immunoadsorbents, a step was included to fully resuspend the beads. After trypsin digestion, 1 µg (5 µg DQLL) of each anti-peptide antibody was added to the samples, followed by a 1 hr incubation step at 1400 rpm. Beads were washed in triplicate using wash buffer, followed by peptide elution in 50 µL of 0.5% FA in Water + 0.03% CHAPS ^26^.

### LC-MS analysis

#### Immuno-MRM

LC separation was performed on an ACQUITY UPLC I-Class FTN system equipped with a Binary Solvent Manager ^19^. Ten microliter of each sample was injected onto an ACQUITY PREMIER Peptide BEH C18 column (2.1 mm × 30 mm, 1.7 μm, 300 Å) (Waters Corporation, Wilmslow, UK). Peptide separation was achieved using gradient elution with mobile phase A consisting of LC–MS-grade deionized water with 0.1% (v/v) FA, and mobile phase B consisting of LC–MS-grade acetonitrile with 0.1% (v/v) FA. Gradient elution was performed at 0.8 mL/min with initial inlet conditions at 5 % B, increasing to 15 % B from 0.15 to 0.35 min and at a steady state for 0.25 min. Subsequently, over 0.4 min, gradient B was increased to 25%, followed by a column wash at 90 % B for 0.25 min and returning to initial conditions at 5 % B. The total run time was 1.8 min. For the 8-min run, gradient elution was performed at 0.6 mL/min with initial inlet conditions at 5 % B, increasing to 33 % B over 5.5 min, followed by a column wash at 90 % B for 1.4 min and a return to initial conditions at 5 % B. The total run time was 8 min. The MS source conditions of Xevo™ TQ-XS (Waters Corporation) conditions were: capillary voltage 0.5 kV, source temperature 150 °C, desolvation temperature 600 °C, cone gas flow 150 L/h, and desolvation gas flow 1000 L/h. The instrument was calibrated at unit MS1 and MS2 mass resolution. Endogenous and the stable isotope labelled peptides were detected using MRM acquisition with experimental details described in Supplementary Table 1.

### Untargeted Acquisition

An ACQUITY M-Class UPLC System (Waters Corporation) was used and operated in trap-elute LC mode with the SCIEX ZenoTOF 7600 mass spectrometer. Mobile phases consisted of 0.1% FA in water (A) and 0.1% FA in acetonitrile (B). The samples were loaded at 10 µL/min with 98.5% mobile phase A and trapped on a TriArt C18 guard column (YMC) (500 µm x 5 mm, 3 µm) for 1.5 min. Microflow separations were done using a YMC Triart C18 (300 µm x 15 cm, 3 µm) at a flow rate of 5 µL/min, using a non-linear LC gradient described in Supplementary Table 2.

An OptiFlow Turbo V ion source was used with the microflow probe (1-10 µL/min electrode) for the capillary flow experiments, with source parameters as follows: GS1 = 20 psi, GS2 = 55 psi, Curtain Gas = 45 psi, CAD gas = 7 psi and Ionspray voltage = 4500 V. For DDA (a cycle time of 0.55 s), MS1 spectra were collected between 400–900 *m/z* for 100 ms. The 45 most intense precursors ions with charge states 2–4 that exceeded 300 counts/s were selected for fragmentation, and the corresponding fragmentation MS2 spectra collected between 140–1800 m/z for 10 ms. After the fragmentation event, the precursor ions were dynamically excluded from reselection for 4 s. Zeno SWATH DIA experiments used 85 variable windows (Supplementary Table 3), spanning the TOF MS mass range 400-900 *m/z* and MS/MS mass range 140-1800 *m/z*, with Zeno trap pulsing turned on, with MS/MS accumulation times of 13 ms.

### Data processing

Skyline-Daily (version 24.1.1.398) was used to process the raw LC–MS data using a template file containing the target peptides of Influenza A, Influenza B, RSV and SARS-CoV-2. Peak integration boundaries were automatically set on the heavy standard and manually reviewed before exporting a report containing the peptide-modified sequence, retention time and heavy normalized area. Metaboanalyst (v6.0) was used for ROC analysis ^30^.

## RESULTS

We and others have demonstrated that LC-MS can reliably detect tryptic peptides unique to SARS-CoV-2 proteins in nasopharyngeal (NP) swabs. However, matrix interferences, originating primarily from preservation media like Universal Transport Medium (UTM), adversely affected sensitivity, challenging the ability to achieve relevant detection thresholds ^11^. To overcome this obstacle, an affinity purification step was implemented using anti-peptide antibodies, which mitigated matrix effects across various transport media and enhanced assay performance in diverse biological matrices, including saliva and plasma ^18–20,31^.

Given that the nucleocapsid protein (N) is the most abundant viral protein per SARS-CoV-2 virion, peptides derived from N were prioritized. Candidate peptides were systematically evaluated for sensitivity and linear dynamic range using both untargeted and targeted LC-MS experiments. Through this process, peptide AYNVTQAFGR (AYN) emerged as an optimal surrogate candidate due to its robust MRM response and resilience against known SARS-CoV-2 mutations—as confirmed by sequences submitted to the Global Initiative on Sharing All Influenza Data (GISAID) database (https://gisaid.org/) ^32^. Although peptide ADETQALPQR (ADE) exhibited mutations in several variants, including Delta, it was retained as a secondary target to ensure robust detection across diverse viral strains ^19^. A similar strategy was applied to identify optimal surrogate peptides for respiratory syncytial virus (RSV), influenza A (IAV), and influenza B (IBV). Each candidate peptide was evaluated based on MRM response, and only those with greater than 99% sequence stability across all human strains reported in GISAID EpiFlu™ and EpiRSV™ were selected. The peptides were then incorporated, alongside the SARS-CoV-2 targets, into a second generation QconCAT heavy labelled internal standard (PolyQuant, Bad Abbach, Germany), which was used here throughout the assay development process. This approach enhanced reproducibility and accuracy but also ensured traceability across laboratories, aligning with ISO 17511:2020 standards ^33^. Affinity-purified rabbit polyclonal antibodies were then generated against two peptides per virus (**Table 1**) and incorporated into an iMRM method designed to quantify these surrogate peptides in tryptic digests of NP swabs stored in UTM. Proprietary methodologies were employed by SISCAPA for the production of the polyclonal antibodies *i.e.*, immunization, antibody screening, and production. All six polyclonal antibodies were assessed in a sequential capture experiment with 0–5 µg of each antibody incubated in 10 µL of trypsin-digested UTM spiked with 1 pmol of stable isotope-labeled “heavy” peptides. Recovery was calculated by comparing amounts recovered after one purification step to the total after two consecutive enrichment steps (Supplementary Figure 2). Antibodies against VII (RSV), GVF (IAV), and VLS (IBV) performed best with a recovery of 70–100%, while MVL (IAV) and SMV (IBV) showed ∼40% recovery, and DQL (RSV) was even unsuccessful.

**Table 1:**
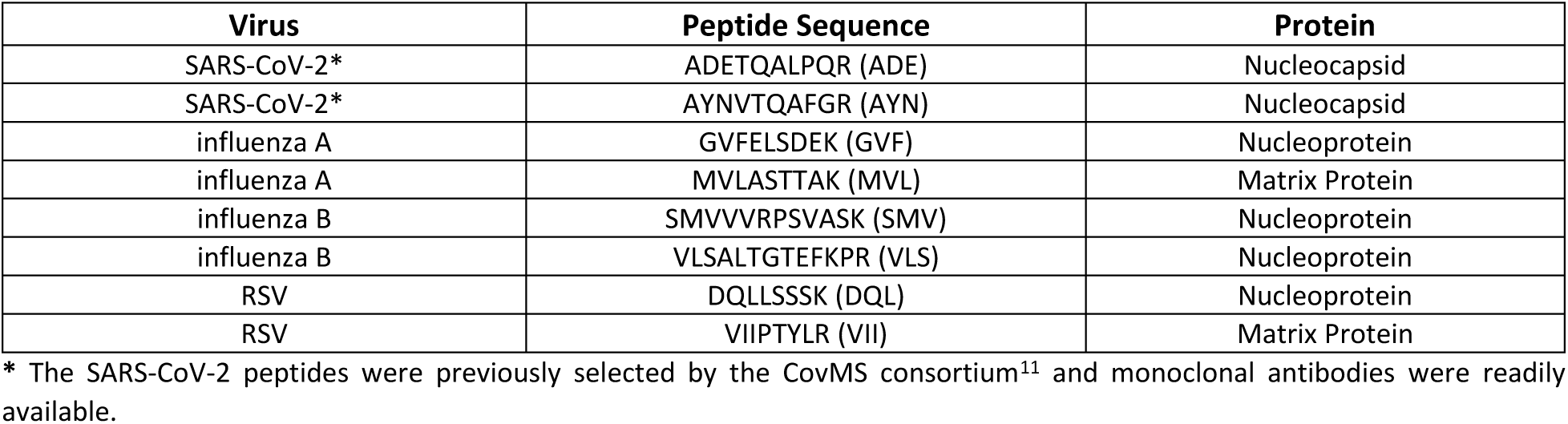
Proteotypic peptide targets selected from SARS-CoV-2, RSV, IAV, and IBV.

### Analytical Validation

Starting with the original 2– and 8-min LC gradients applied by the CovMS consortium ^11^, we added the new peptide targets for IAV, IBV, and RSV following collision energy optimization of the selected fragment ions. Surprisingly, these additional peptides were nicely distributed across both gradients, which allowed us to optimize scan times and thus enhance sensitivity. This optimized approach seamlessly integrated with the automated Andrew+™ automated CoV2MS method, supporting a robust, multiplexed LC-MS-based platform for viral surveillance (**Figure 1**). Moreover, it readily affords the possible inclusion of additional target peptides, enabling the detection of other common respiratory infections such as human metapneumovirus or parainfluenza viruses in the future. Chromatographic separation of the target peptides during a 2-min injection-to-injection run was achieved, with peptides ADE (SARS-CoV-2), DQL (RSV), MVL (IAV), SMV (IBV), AYN (SARS-CoV-2), GVF (IAV), VLS (IBV), and VII (RSV) eluting at retention times of 0.47, 0.49, 0.53, 0.57, 0.72, 0.76, 0.90, and 1.05 min, respectively. No significant interfering peaks were observed at the retention times of the endogenous peptides or their corresponding stable isotope-labeled standards (SILs).

**Figure 1.**
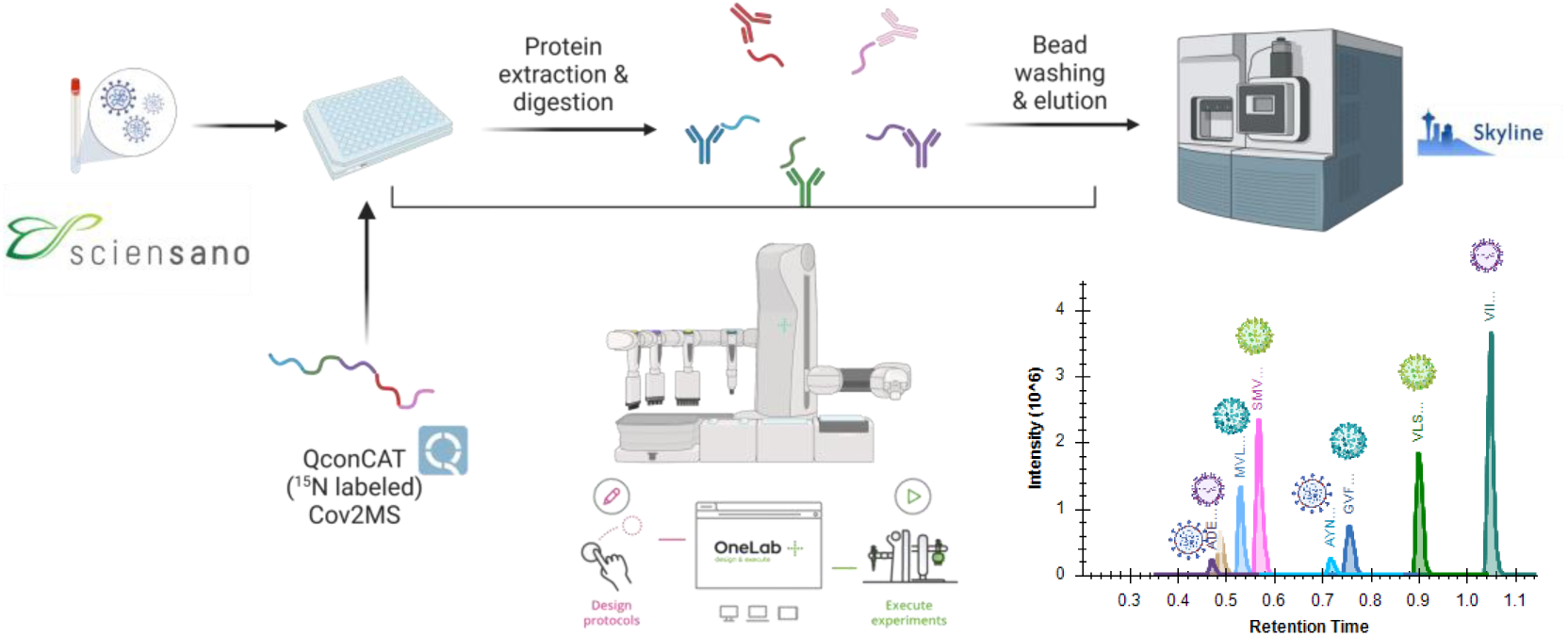
Workflow of the Viral Respiratory iMRM Winterplex Method. Schematic overview of the iMRM workflow for the quantification of SARS-CoV-2, influenza A, influenza B, and RSV viral loads from clinical nasopharyngeal swabs, obtained through Sciensano, Belgians national reference center for respiratory pathogens. Proteins are extracted, digested, and enriched using anti-peptide antibodies, followed by extensive washing and acidic elution. A QconCAT (^15^N-labeled Cov2MS) internal standard is used for absolute quantification. The enriched peptides were analyzed using a 2-min LC-MS method with data processing performed using Skyline software. The chromatogram (bottom right) illustrates the separation and detection of different viral peptides across the 2-min LC run.

The linearity of the method was assessed according to the CLSI EP06-A guideline, using 10 replicates of a mixture of the synthetic peptides (0-1000 amol/µL) spiked into digested QconCAT internal standard (1.5 fmol/µL) and measured with both LC gradients. Overall, both gradients demonstrated similar performance in terms of linearity, and we therefore opted to continue with the 2-min method (**Figure 2a**), since it provides throughputs in the range of 500 measured samples per instrument per day. These replicate injections also demonstrated reproducible retention times with residual standard deviation (RSD) of ≤ 1%. The calibration curves for almost all peptides exhibited a linear response across the concentration range 7.8 – 1000 amol/µL with a correlation coefficient (R²) of ≈0.99, except for ADE. For this specific peptide, linearity was obtained in the 125 – 1000 amol/µL concentration range, with an R² of >0.999. These findings are consistent with previously reported results.

**Figure 2.**
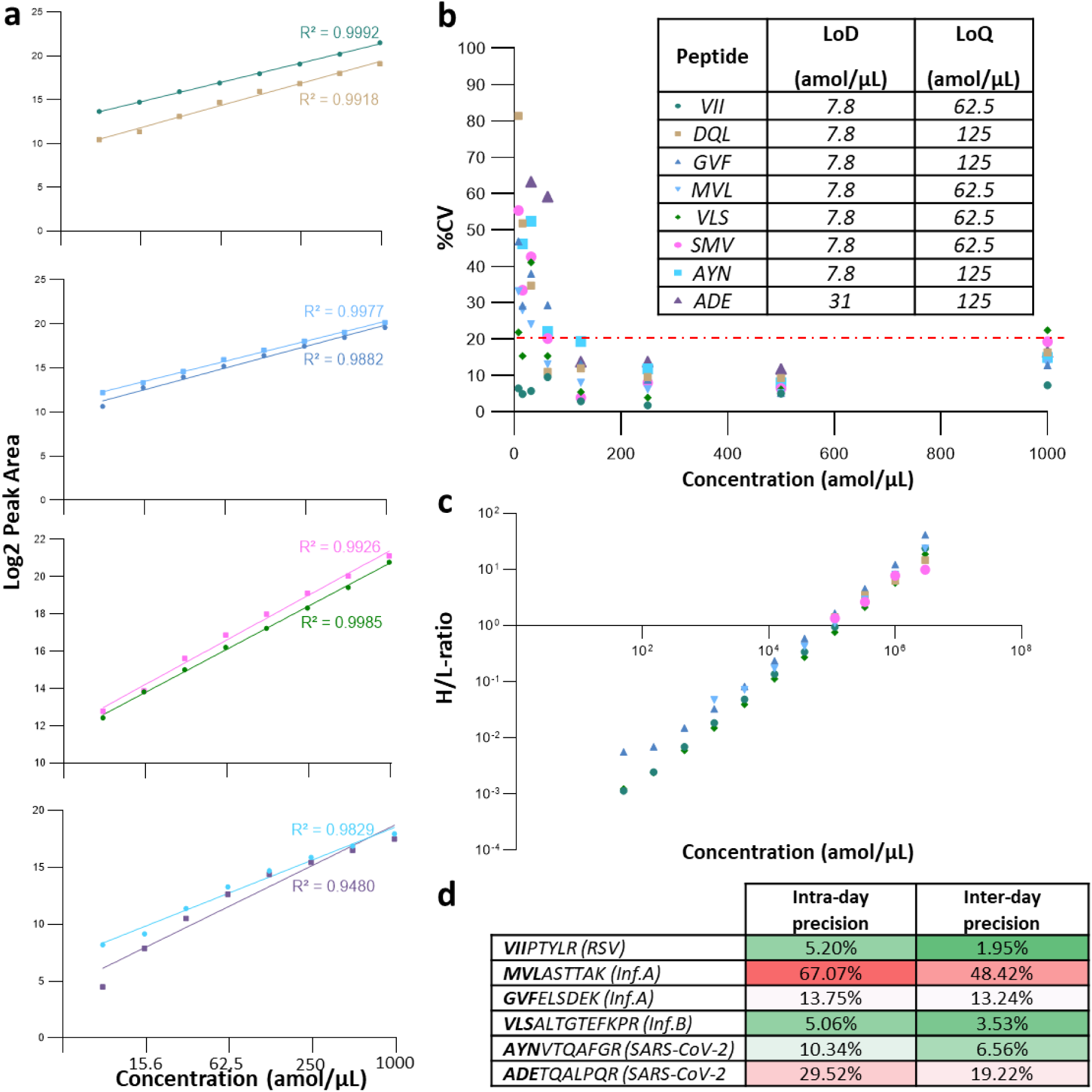
Winterplex iMRM method evaluation. [a] A dilution series of the synthetic peptide targets for each virus (0 – 1000 amol/µL), in digested QconCAT (1.5 fmol/µL) was measured 10 times after which the linearity and the corresponding correlation coefficient for each peptide were determined. The blank sample was removed from data analysis, as the signal for most peptides was zero. [b] Percent coefficient of variation (%CV) plot, wherein each colored symbol represents the %CV of the log2 peak area for each target peptide. The red dashed line denotes an acceptance threshold of 20 %CV. Higher %CV values at lower concentrations suggest increased analytical variability in that range. The inset displays the LoD and LoQ values obtained for each target peptide. [c] Linearity of the H/L-ratio across a dilution series of synthetic stable-isotope labelled peptides of Influenza A &B and RSV in digested UTM. [d] Evaluation of the intra– and inter-day precision (n = 5) for all targets using the heavy spiked-in Polyquant standard.

Furthermore, the coefficient of variation (%CV) (**Figure 2b**) and accuracy were also calculated from the ten replicate measurements to determine Limit of Detection (LoD) and Limit of Quantification (LoQ) in accordance with the CLSI EP17-A2 guideline. The LoD represents the lowest concentration at which an analyte can be reliably distinguished from background noise in a blank sample. In contrast, the LoQ is defined as the lowest concentration at which the analyte can be quantified with acceptable precision and accuracy, meeting the criteria of a %CV ≤ 20% and a measured concentration within 80–120% of the expected value. For peptide AYN, the LoQ was determined to be 125 amol/µL, with an LoD of 7.8 amol/µL. Similarly, peptide ADE had an LoQ of 125 amol/µL, though its LoD was slightly higher at 31 amol/µL. Peptide GVF also exhibited a LoQ of 125 amol/µL, with a LoD of 7.8 amol/µL. In contrast, peptides MVL, VLS, SMV, DQL, and VII demonstrated lower LoQ values of 62.5 amol/µL, while maintaining the same LoD of 7.8 amol/µL (**Figure 2b, inset**). Furthermore, a standard curve (50 – 3×10⁶ amol/µL) was generated by titrating the synthetic stable isotope-labeled (SIL) peptides in UTM, spiked with a constant 1,000 fmol of synthetic “light” peptide in all samples. The heavy-to-light peptide ratio was used, resulting in LoD and LoQ values that were similar to or slightly higher for VLS, GVF, and VII, highlighting the antibodies’ efficient enrichment (**Figure 2c**). In contrast, the limits for DQL, SMV, and MVL were significantly higher, reflecting the weaker performance of their respective polyclonal antibodies.

Besides the dilution series acquired in pure elution buffer and UTM to benchmark the Winterplex iMRM method, a dilution series was included before and after each sample plate. Additionally, immediately after each sample plate and dilution series, a blank sample was measured. This comprehensive setup allowed for the assessment of instrument sensitivity and carryover of the iMRM method across sample batches, following the guidelines of CLSI EP10. No explicit carryover was observed for any of the peptides analyzed, as evidenced by the absence of residual signals in blank samples following the highest calibrator measurement (1000 amol/µL), underscoring the assay’s robustness and ensuring that potential carryover does not compromise the accuracy and reliability of the results (Supplementary Figure 3). Notably, we have previously observed such behavior for the KQQTVT peptide of SARS-CoV-2, further supporting the reliability of our approach ^19^.

The precision of the method was assessed at two concentrations of synthetic peptide spikes, namely 2000 amol/µL and 400 amol/µL, added to confirmed negative patient samples in UTM, with five replicates tested over five consecutive days. Due to limited antibody availability, precision could not be assessed for peptides DQL (RSV) and SMV (IBV). At all three concentrations, both inter– and intra-day precision exceeded 20%. However, further analysis revealed that there was no issue with the heavy Polyquant standard (1.5 fmol/µL), suggesting that the observed variability likely stemmed from inconsistent spiking of synthetic peptides rather than from variability introduced by affinity enrichment or iMRM measurement. Interestingly, when reproducibility was assessed solely based on the heavy Polyquant peptides, intra– and inter-day precision remained below 20% CV for one peptide per virus (**Figure 2d**), despite the expected variability introduced by the protein digestion step. Notably, the MVL peptide exhibited significantly higher variability, likely due to lower binding efficiency as described before, while the precision of the ADE peptide was probably affected by the use of only half the recommended amount of monoclonal antibodies due to limited supply.

### Measurement Response

The variability in retention times and sensitivity of the Xevo TQ-XS triple quadrupole (QqQ) instrument was assessed by monitoring a mixture of SIL target peptides (0.5 fmol/µL). This QC sample was interspersed throughout the sample batch, with data automatically imported into Skyline Daily via the AutoQC loader and uploaded to Panorama QC for systematic tracking ^34^. Since these peptides were labeled differently from the heavy-labeled QconCAT, no interference with the analyzed target peptides was observed. Retention times for all peptides remained stable throughout the entire batch, spanning approximately nine days. However, MRM detection sensitivity declined by nearly 50% (Supplementary Figure 4), suggesting some form of charging or buildup of contamination, despite extensive sample purification with polyclonal antibodies and three consecutive washes to remove unbound peptides and other unwanted contaminants, such as salts. An additional washing step could be implemented in future workflows to further minimize background contamination while preserving absolute detection sensitivity. Moreover, recent instrument advancements aimed at reducing ion charging are expected to further enhance overall robustness ^35^.

Despite the decline in sensitivity, the use of a heavy-labeled QconCAT mitigated its impact on quantitative accuracy. This is particularly critical for the future clinical implementation of the Winterplex method, where analytical precision and consistency are essential for maintaining diagnostic reliability.

### NP Patient Samples

A total of 344 residual clinical NP swab samples confirmed positive for IAV (H1N1 or H3N2), IBV (Victoria or Yamagata), RSV (A or B), or SARS-CoV-2 by RT-qPCR were processed using the automated sample preparation workflow (Supplementary Figure 5) and subsequently analyzed using a 2-min Winterplex iMRM method. The cohort comprised 21 SARS-CoV-2, 58 IAV H3N2, 47 IAV H1N1, 108 IBV, and 110 RSV samples with Ct values ranging from 12.52 to 39 (Supplementary Figure 6). Due to limited anti-peptide antibody availability, a subset of 87 samples per virus type (excluding SARS-CoV-2) was prepared separately in a 96-well plate and enriched only for the corresponding viral peptides.

During method development, untargeted data-dependent acquisition (DDA) was performed on a selection of samples following immunopurification to validate the enrichment efficiency of the polyclonal antibodies. Despite the extensive background signals typically encountered in NP swabs stored in UTM, fewer than 50 peptide sequences were detected in immunopurified IAV samples, including the affinity enriched target peptides MVL and GVF. This low number of identified peptides further confirms the high efficiency of the enrichment process, consistent with what has been previously reported. Notably, the miscleaved peptide GRGVFELSDEK, unique to the H3N2 subtype, was also identified and subsequently added to the iMRM method to enable IAV subtyping. As illustrated in **Figure 3a**, the miscleaved GRGVFELSDEK peptide effectively differentiates H3N2 (green) from H1N1 (red) samples, achieving 95% specificity. Furthermore, the linear inverse correlation between LC-MS signals and RT-qPCR Ct values highlight the reliability of GRGVFELSDEK for both subtyping and determining the viral load of Influenza A strains.

**Figure 3.**
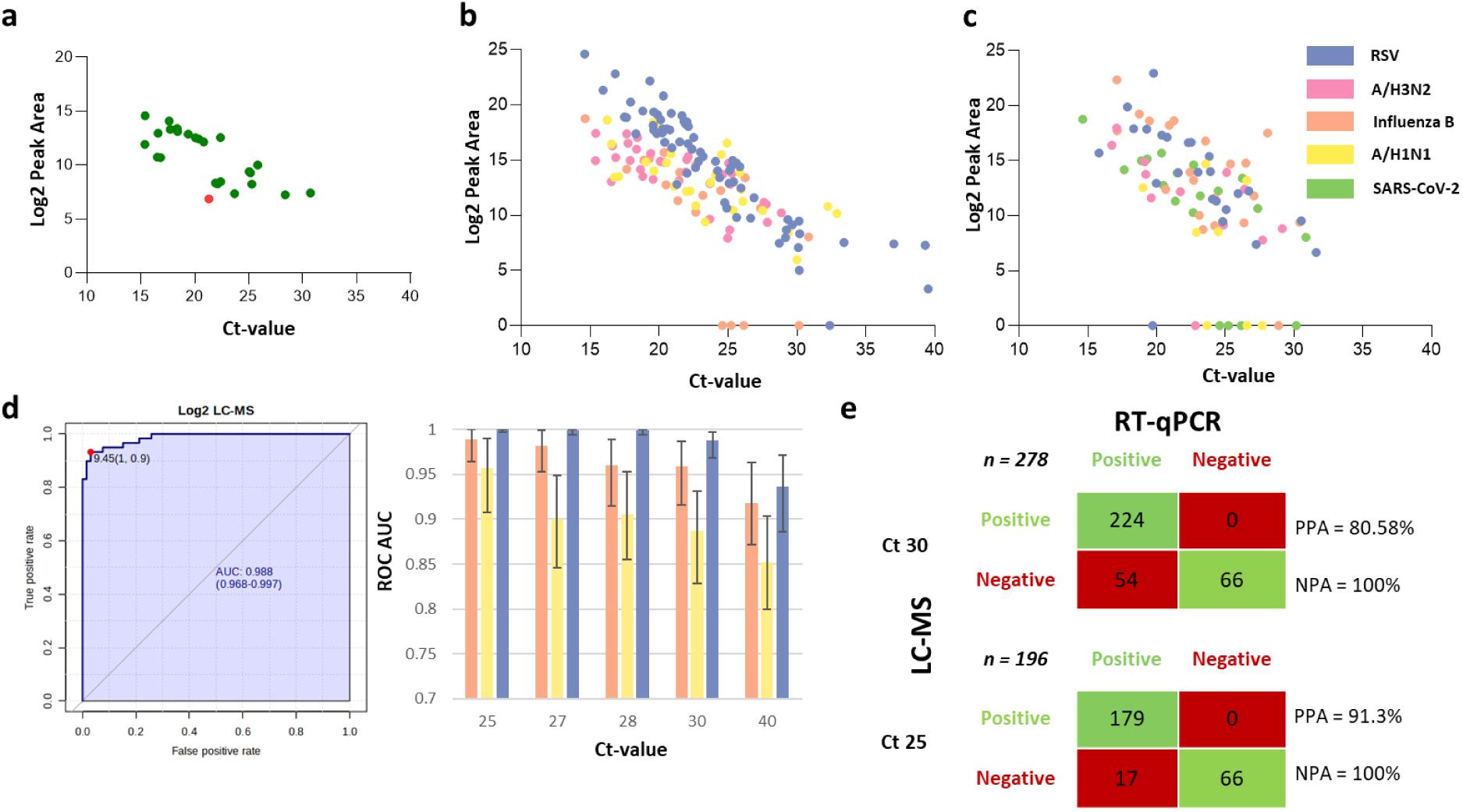
Correlation between LC-MS (log₂ summed peak area) and RT-qPCR cycle threshold (Ct) in nasopharyngeal swab samples positive for influenza A, influenza B, RSV or SARS-CoV-2. [a] All H3N2 (green) and H1N1 (red) samples with measurable signal for the GRGVFELSDEK peptide were visualized to demonstrate that Influenza A subtyping can be achieved using this target peptide with 95% specificity [b] Three separate 96-well plates, each containing patient samples for a single respiratory virus (RSV, IAV, and IBV), were individually processed using peptide affinity purification to the respective virus. [c] A single 96-well plate containing a mixed cohort of patient samples positive for RSV, IAV, IBV, or SARS-CoV-2 (confirmed by RT-qPCR) was prepared using affinity enrichment for all Winterplex targets. For correlation analysis, the log₂ peak area of a single representative peptide per virus was used. [d] ROC curve with true positives defined by RT-qPCR. ROC Area Under the Curve (AUC) for each virus at different Ct values. Above Ct 30, a noticeable drop-off to an AUC of < 0.95 can be seen, suggesting that from here on, both diagnostic tests start to disagree. Error bars indicate the 95% confidence interval (CI). [e] LC-MS vs. RT-qPCR contingency matrix, used to calculate the Positive Percent Agreement (PPA) and Negative Percent Agreement (NPA) between both detection methods at Ct-values 30 and 25.

**Figure 3b** illustrates the correlation between the log₂ summed peak area obtained via LC-MS and the RT-qPCR cycle threshold (Ct) values for three respiratory viruses: IAV subtypes H3N2 and H1N1, IBV, and RSV. For IAV, the log₂ summed peak area of GVF was used, as MVL was only detectable in a small subset of high-viral-load samples. Among the three monitored light transitions for GVFELSDEK, the y7++ transition was excluded from quantitative analysis due to significant interference in both positive and negative samples. Similarly, correlation with Ct-values for IBV and RSV was assessed using only the log₂ peak area obtained for VLS and VII, respectively, due to the poor performance of the other polyclonal antibodies (see above). Any sample with a heavy peptide peak area below 10% of the average heavy peptide signal (<5%) was also excluded from further analysis, as this likely indicated poor digestion. Notably, three IBV samples with low Ct-values (Ct < 30) exhibited no detectable signal, as illustrated by the orange dots aligned along the x-axis. To investigate whether this absence was due to a mutation in the VLS peptide, we analyzed the flow-through (FT) using untargeted acquisition methods after antibody incubation in order to assess the presence of a mutated version of the target peptide, as described earlier ^19^. Although no known mutated versions of the VLS peptide were detected in the FT of the samples, several other IBV-related peptides were identified in the FT of one of the samples (255525), including six from the nucleoprotein and one each from the matrix and two non-structural proteins, confirming that this patient was indeed IBV positive. Reanalyzing the enriched sample using data-independent acquisition (DIA), did not surface the endogenous or a known mutated version of the VLS peptide either.

Next, a single 96-well plate containing a mixed cohort of RSV, IAV, IBV, and SARS-CoV-2 patient samples was prepared using all Winterplex anti-peptide antibodies, including the less performant ones, to assess the effect of multiplexing. For both designs, LC-MS quantification showed high concordance with the Ct values obtained via RT-qPCR, with Pearson correlation coefficients ≥ 0.7 across all virus groups. The observed linear relationship (Figure 3c) demonstrates the increased LC-MS signal (log₂ summed peak area) corresponding to lower Ct values, indicative of higher viral load. Notably, one sample (255207) in the mixed plate was initially classified by RT-qPCR positive for Influenza A only. However, analysis of the same sample using the Winterplex iMRM method revealed clear signals for both Influenza A and SARS-CoV-2, indicating a dual infection that was subsequently confirmed by RT-qPCR (Supplementary Figure 7a). In fact, once upscaled, multiplexed viral detection by iMRM can be done at very little extra cost and no extra effort, underscoring the added resolution that the Winterplex iMRM can provide in detecting co-infections that might be overlooked or misclassified by single plex methodologies. Furthermore, one sample (255598) was either misclassified as influenza B by RT-qPCR or an error occurred during LIMS registration, because it tested positive for SARS-CoV-2 with iMRM (Supplementary Figure 7b).

**Figure 3d** shows an exemplary Receiver Operating Characteristic Area Under the Curve (ROC AUC) and the confidence intervals obtained at different RT-qPCR thresholds for the individual sample plates. All three newly included respiratory viruses exhibited a perfect agreement (AUC ≥ 0.95) at Ct 25, however at lower viral loads, a noticeable drop-off below an AUC of 0.95 is seen, suggesting that from there on, LC-MS starts to disagree slightly with RT-qPCR. Notably, detection of VLS (IBV) and especially VII (RSV), was less affected by higher Ct-values, which might imply better performance of their respective anti-peptide antibodies. In contrast, the GVF (IAV) signal was weaker, which may reflect the lower efficiency of its polyclonal antibody. It’s worth noting that only 1 µg of the GVF (IAV) antibody was used, while the SISCAPA recommended amount was 2 µg (Supplementary Data). Still, as discussed previously ^19^, apart from differences in Ct distribution of the patient populations for each virus, with the IAV sample cohort displaying an overall higher median Ct-value compared to the other respiratory viruses, many more reasons can underly a discrepancy between LC-MS and RT-qPCR, with no specific reason to prefer one over the other. In conclusion, while this comparison provides an analytical framework of assay performance, it does not validate the performance of the Winterplex iMRM assay on patient samples directly.

A contingency matrix comparing LC-MS results with RT-qPCR Ct-values for all samples is presented in Figure 3e. Positive Percent Agreement (PPA) and Negative Percent Agreement (NPA) were calculated for each virus at Ct thresholds of ≤ 30 and ≤ 25. Samples in the mixed plate that tested positive for one virus were used as negative controls for the other viruses. PPA increased from 80% at Ct 30 to 91.3% at Ct 25, demonstrating strong concordance in detecting true positives across influenza A, influenza B, and RSV. Meanwhile, NPA remained at 100%, indicating that LC-MS reported no false positives relative to the RT-qPCR reference standard.

### Avian flu

During peptide selection, no distinction was made between human and non-human influenza A strains, and only sequence conservation across human strains was considered, without accounting for potential overlap with non-human strains. To evaluate this, a highly concentrated, β-propiolactone (BPL)-inactivated avian influenza isolate (A/Gallus_gallus/Belgium/00548_0001/2023) was processed similarly to the NP swab samples but without peptide immuno-enrichment and using untargeted MS acquisition via data-dependent (DDA) and data-independent (DIA) acquisition. Both target peptides (GVF and MVL) yielded strong signals in both acquisition modes, as demonstrated by the DIA data, presented in Figure 4. Additionally, 80 peptides corresponding to 9 of the 12 known next-generation sequenced proteins uploaded to GISAID were identified, with over 50% mapping to the Nucleoprotein and Matrix protein.

**Figure 4.**
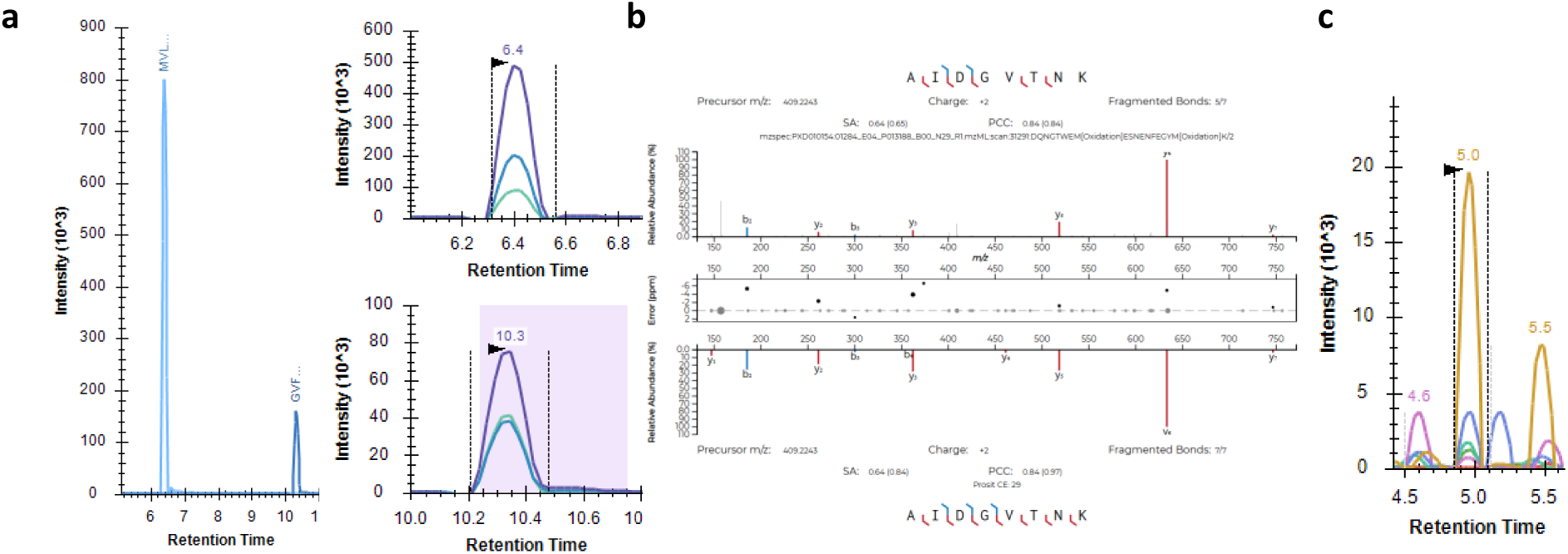
Untargeted DIA analysis of an inactivated avian influenza (H5N1) sample. [a] An exemplary extracted ion chromatogram (XIC) displaying the two influenza A target peptides, MVLASTTAK (top) and GVFELSDEK (bottom). While these peptides enable influenza A detection, they do not allow for subtype differentiation. [b] A peptide specific to Hemagglutinin (AIDGVTNK) was identified as unique for H5N1. The Pearson Correlation Coefficient (PCC) between the measured and Prosit-predicted spectra exceeded 0.8; however, [c] the fragment ion signal intensity was lower compared to influenza A peptides GVF and MVL.

To assess cross-reactivity, peptide immuno-enrichment was performed on a dilution series of the inactivated avian flu culture using the GVF and MVL polyclonal antibodies. As expected, both peptides were successfully enriched, even in the lowest tested concentration, further supporting their potential for broad influenza A surveillance in both human and avian populations. However, to enhance diagnostic specificity for avian influenza (H5N1), future efforts should focus on developing antibodies against peptides uniquely derived from avian strains. Therefore, we leveraged both DDA and DIA data from the H5N1 isolate, to identify AIDGVTNK as a candidate Hemagglutinin-derived peptide, which is notably absent in human influenza strains. This selection reflects the well-documented sequence diversity of Hemagglutinin and Neuraminidase, two surface glycoproteins critical for host cell attachment, pathogenicity, and viral progeny release ^36,37^. Nevertheless, given the accelerated evolutionary rate of avian influenza strains compared to their human counterparts, identifying a peptide that remains conserved over time remains a significant challenge.

The correct annotation of this peptide was further validated by calculating the Pearson Correlation Coefficient (PCC) between the experimentally measured MS2 spectrum and the theoretical spectrum predicted by Prosit, a machine-learning-based fragment ion intensity predictor ^38,39^. Indeed, this exceeded 0.8, as shown in Figure 4b, in the process confirming the y6 ion as a promising transition for a future assay. Again, immuno-enrichment will be necessary, as exemplified by the increased number of interfering transitions in the DIA data (Figure 4c) and the lower overall fragment ion intensity—compared to GVFELSDEK and MVLASTTAK.

## DISCUSSION

RT-qPCR remains the gold standard for viral detection due to its high sensitivity and the lack of a specific demand for quantitative accuracy in diagnostic testing. However, the SARS-CoV-2 pandemic highlighted the limitations of relying on a single technology, which can undermine societal resilience ^40^. From day one, numerous mass spectrometers at universities and biotech companies worldwide remained idle, despite being extremely sensitive and the most accurate analytical tool available for biomolecular quantification ^41^. What was missing was a well-defined, SOP-documented, and validated diagnostic assay for the quantification of respiratory viruses. Over the past five years, we have developed such an assay and demonstrated that peptide immuno-enrichment reagents are crucial for enhancing sensitivity, throughput, and robustness. This manuscript presents the final key optimization, namely multiplexing. Beyond showcasing this capability, we view this work as a call to action, urging policymakers and public health agencies call to invest in a comprehensive pathogen detection portfolio. Developing reagents for all potential viral threats as part of pandemic preparedness would provide society with a critical buffer in terms of testing capabilities, significantly improving our ability to contain future viral outbreaks ^42^.

LC-MS, in combination with peptide immuno-enrichment, offers a scalable and multiplexed platform for respiratory virus detection. By targeting at least one proteotypic peptide per virus and applying stringent validation criteria, we achieved high concordance with RT-qPCR results, particularly at Ct values below 30. However, concordance at higher Ct values decreases, potentially reflecting differential persistence of these two biomolecules in disease progression. RT-qPCR detects both infectious and non-infectious viral nucleic acids, whereas LC-MS identifies specific viral peptides, potentially offering greater specificity for detecting active infections ^19^. While we hypothesize that LC-MS preferentially detects active infections, this requires further validation. Regardless, it is clear that LC-MS generally exhibits lower sensitivity than RT-qPCR, which may limit its ability to detect low viral loads in scenarios where patient isolation is critical. These inherent differences in detection mechanisms highlight the trade-offs between sensitivity and specificity when considering LC-MS as a complementary or alternative diagnostic approach to RT-qPCR.

Key advantages of LC-MS include its ability to support large-scale testing and its potential for almost unlimited multiplexing. The Winterplex iMRM method demonstrated consistent reproducibility, and by leveraging immuno-enrichment, it effectively mitigated matrix interferences, making it compatible with practically any biological sample. Nevertheless, there are several limitations to the present study that highlight critical areas requiring further investment. Not all clinical samples could be tested for all viral targets due to a limited supply of polyclonal antibodies, highlighting the urgent need for scalable and sustainable production of high-quality immunoreagents. Antibody development remains a critical determinant of assay performance, as evidenced by the lower sensitivity observed with the polyclonal antibodies for MVL, DQL and SMV. Future iterations of the Winterplex platform should incorporate high-affinity monoclonal antibodies targeting at least two proteotypic peptides per virus to improve assay robustness and lower detection thresholds. In addition, the method can be continuously expanded by integrating new targets to support viral subtyping and the detection of other relevant respiratory pathogens, such as human parainfluenza viruses (hPIVs) and human metapneumovirus (hMPV), both of which have shown increased incidence in the post-COVID-19 era^43,44^.

Additionally, discrepancies between RT-qPCR and LC-MS results remain unresolved. These are likely attributable to two key factors: (1) a time gap between the two analytical workflows, potentially leading to differential degradation of RNA and protein, and (2) the lack of a standardized calibrator suitable for simultaneous use across both platforms. The development of such a dual-use standard will be essential to ensure analytical alignment and cross-platform comparability. These limitations are not technological dead ends, but rather clear indicators of where strategic investment is required. The COVID-19 pandemic demonstrated that, with sufficient resources and coordinated efforts, complex analytical technologies can be rapidly scaled and deployed.

Furthermore, while this study primarily focused on method validation, the integration of LC-MS into routine diagnostic workflows will require further optimization, particularly for high-throughput applications. The use of Acoustic Ejection Mass Spectrometry (AEMS) has already been explored as a mean to increase processing speed and, with the ability to handle up to 6,500 samples per day, holds significant promise for pandemic preparedness and large-scale surveillance efforts ^45^.

Overall, this study demonstrates the feasibility of LC-MS–based multiplexed detection of respiratory viruses as a complementary and potentially orthogonal approach to RT-qPCR. With ongoing advancements in automation and high-throughput workflows, LC-MS has strong potential to enhance diagnostic capacity for routine surveillance and outbreak response. Importantly, our findings also underscore the critical need to invest in pandemic preparedness between outbreaks, particularly in the development and validation of high-performance monoclonal antibodies, to ensure rapid deployment and scalable diagnostics when the next pandemic emerges.

## CONCLUSION

The resulting Winterplex targeting SARS-CoV-2, RSV, influenza A, and influenza B, has been extensively validated, confirming its suitability for high-throughput viral surveillance. Our findings enhance influenza surveillance capabilities and strengthen preparedness for future outbreaks and pandemics. By improving and expanding reagent development to other viruses of concern during inter-pandemic phases, this assay can be rapidly scaled for widespread deployment when new threats emerge.

## DATA AVAILABILITY

The mass spectrometry proteomics raw data (MRM, DDA and SWATH) files have been deposited to the ProteomeXchange Consortium via the Panorama Public partner repository with dataset identifier PXD064656 (https://panoramaweb.org/winterplex.url). Data can be accessed using the reviewer account details, (Password: nQmwyk3%lZIH?6).

### Potential Conflicts of Interest

The authors declare the following competing financial interest(s): Van Hulle M. and Vissers J.P.C. are employed by Waters Corporation.

### Research Funding

Funding from Research Foundation Flanders (FWO) [1278023N] (B.V.P.) and European Unions’ Horizon Research and Innovation Program [OneLab – No. 101073924].

## Acknowledgements

The authors thank Céline De Sterck, Yuna Stevens and Diana Isabel at the NRC Influenza for their support with sample processing and RT-qPCR testing, and Florian C. Sigloch from PolyQuant GmbH for providing the Cov2MS QconCAT heavy standard.

## AUTHOR CONTRIBUTIONS

B.V.P., J.P.C.V. and M.D. contributed to conceptualization; F.D., S.D., A.P., T.V.M, C.B., K.D.C., S.V.G. provided SARI patient samples. D.D. helped with funding acquisition; M.V.H. and J.P.C.V. helped with the MRM measurements. B.V.P., J.P.C. and M.D. wrote the original draft and all authors have proofread the manuscript.

## Notes

### Author Declarations

Residual nasopharyngeal samples screened for SARS-CoV-2, influenza B, influenza A and RSV by RT-qPCR were obtained from the Central Biobank Platform Sciensano (Ixelles, Belgium), with approval of the University Hospital Ghent ethics committee (ONZ-2023-0500)

