## Supplementary Information for "An LC-immuno-MRM-MS Winterplex Method Development Framework for Respiratory Viral Screening"

Corresponding author:

### Supplementary Figures

MGTKGAIETEPAVKITFGGPSDSTGSNQNGERGQGVPINTNSSPDDQIGYYRRATANKDGIIWVATEGALNTPKDHIGTRNPANNAAIVLQLPQ
GTTLPKAYNVTQAFGRRGPEQTQKKK**ADETQALPQR**QKDKKK**AYETQALPQR**Q**KKQQTVTLLPAADLDDFSK**LIQNSLTIERMVLSAFD
ERRNRYLEEHPSAGKDPKKTGGAQRAMMDQVRESRYDFEKEGYSLVGIDPFKLLQNFRGR**GVFELSDEK**ATNAQRLEDVFAGKNTDLEVKTR
PILSPLTKGILGRKLKREITFHGAKEISLHENR**MVLASTTAK**TIGTHPSSSAGLKAYQKRMGVQMQRFKTEIKKSVYNMVVKSSYVGDEASSKAG
LNDDMERALVDQVIGSR**SMVVVRPSVASK**VVLPISIYAKGAAYEDLR**VLSALTGTEFKPR**SALKLTDIQKALIGASICFLKPKDQERKRRFITE
PLSGMGTTATKKKGLMQVKLGTLCALCEKLAEELQSNIGVLRGEGIAKDVMEVLKLNDTLNK**DQLLSSSK**YTIQRLGGEAGFYHILNNPKNQDL
YDAAKVTSKK**VIIPTYLR**SISVRMEKFAPEFHGEDANNRATKFLESIKGKFTSLVSFKEDPTPSDNPFSKLNSPKIRVYNTVISYIESNRKNNKKTI
KNTLDIHKSITINNPKIVPEPQPKLAAALEHHHHHH

**Supplementary Figure 1. Polyquant Cov2MS Protein Sequence**. Peptides in the Winterplex method are color-coded by virus: SARS-CoV-2 in green, influenza A in yellow, influenza B in purple, and RSV in red. The construct also includes additional peptides for all four viruses; however, these are not part of the Winterplex iMRM panel.

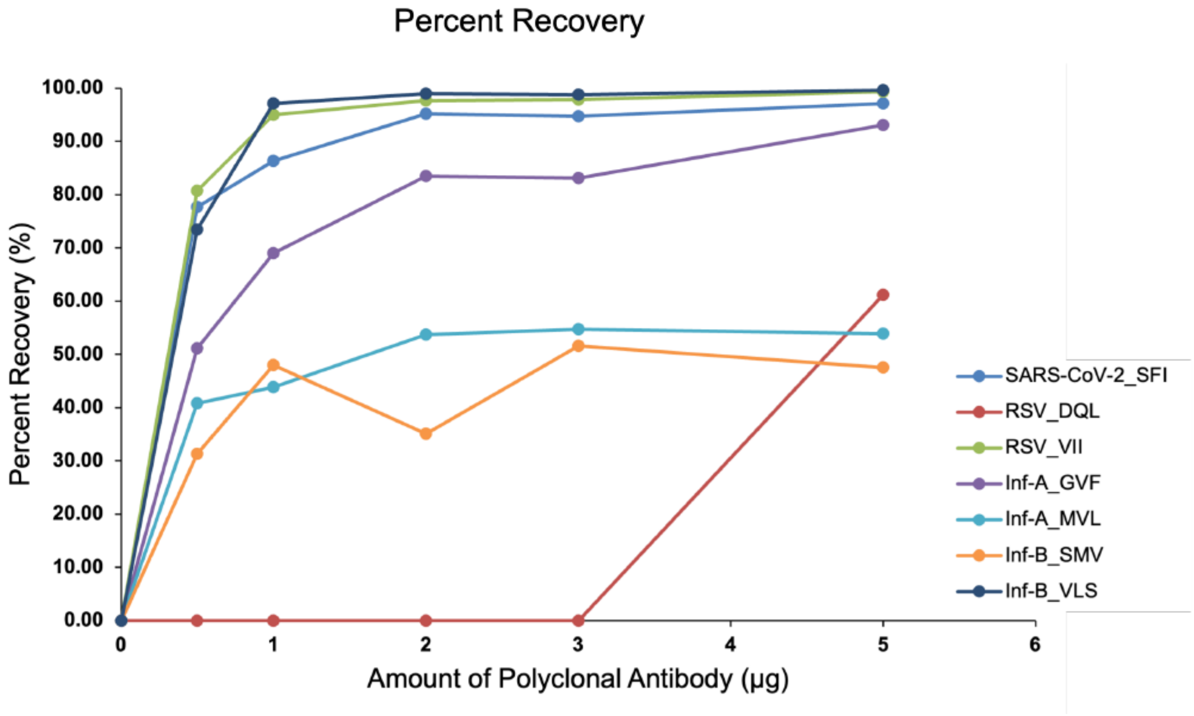

**Supplementary Figure 2. Polyclonal antibody characterization.** Recovery of 1 pmol of stable isotope-labeled (SIL) peptide from trypsin-digested universal transport medium using varying amounts (0-5 µg) of polyclonal antibodies coupled to Dynabeads Protein G.

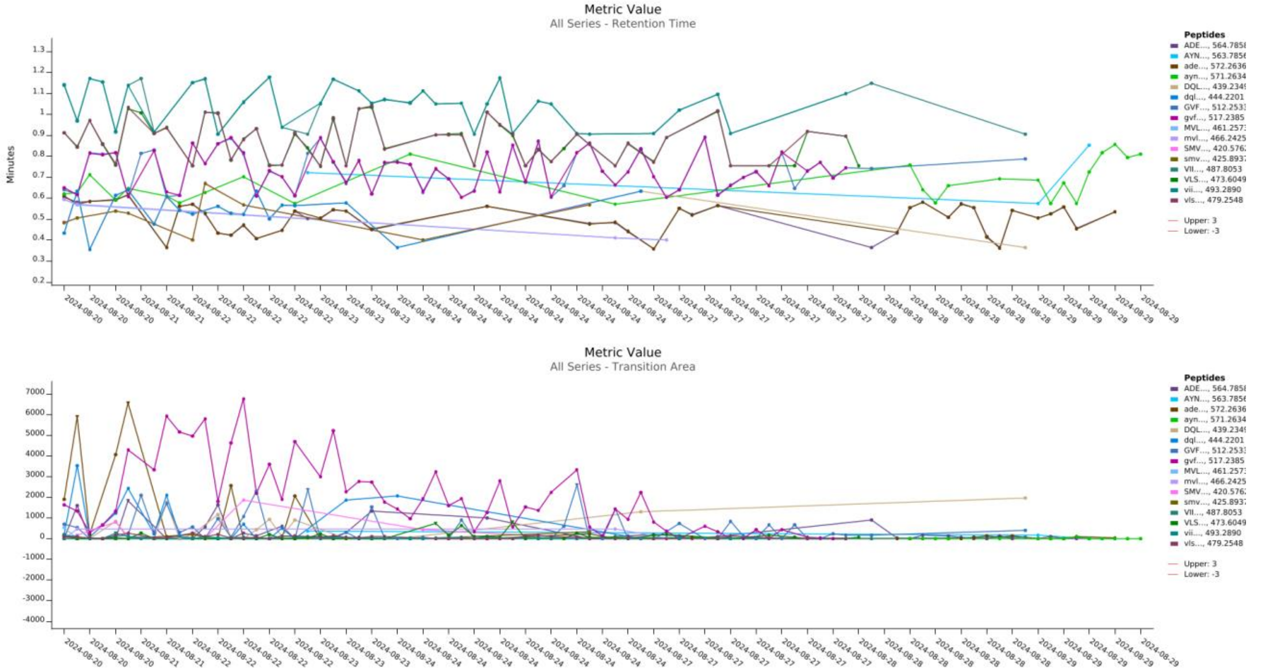

**Supplementary Figure 3. Carry-over assessment.** The carry-over was assessed by running a blank sample after every single full sample plate which also included a dilution series at the beginning and end of a sample plate. [Top] The variability in retention time of both the endogenous and heavy peptides indicates that no distinct signal can be detected for all peptides. [Bottom] This is supported by the very low peak area measured for all individual endogenous peptides.

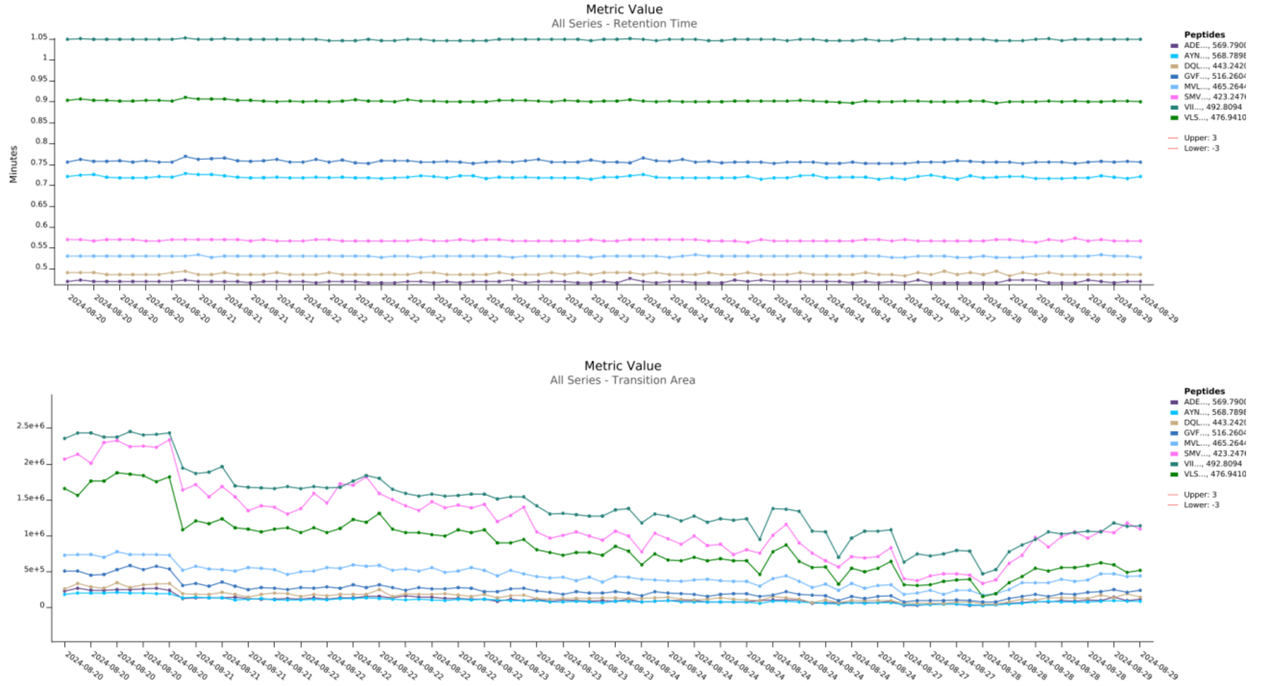

**Supplementary Figure 4. Panorama QC – System performance.** A QC mixture containing the different peptides with a stable isotope label was measured repeatedly throughout the sample batch to monitor the performance of the Triple Quadrupole Xevo TQ-XS instrument. [Top] The retention time (RT) for all peptides remained constant throughout the 9 days of acquisition. [Bottom] The peak area shows a drop over time which could indicate potential contamination or drifting of the instrument calibration.

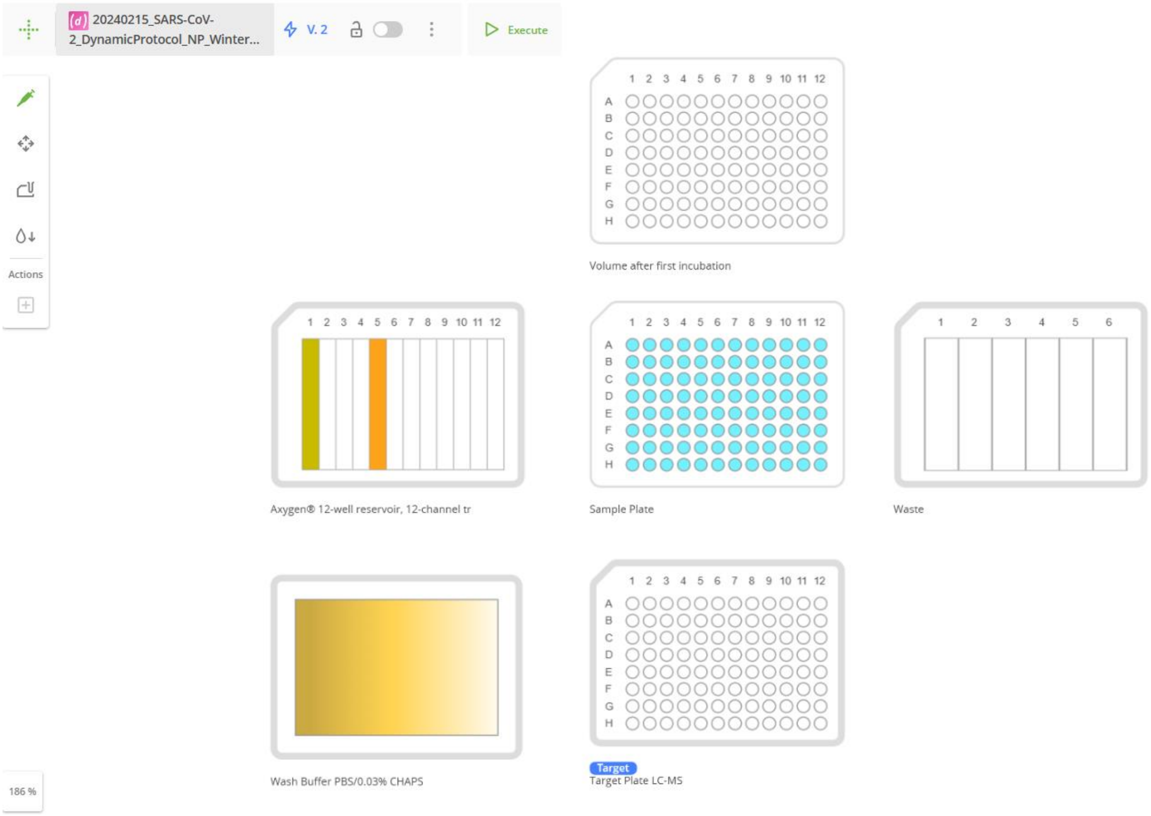

**Supplementary Figure 5.** Deck overview of the Andrew+ protocol for the automated sample preparation of nasopharyngeal swabs for viral respiratory screening.

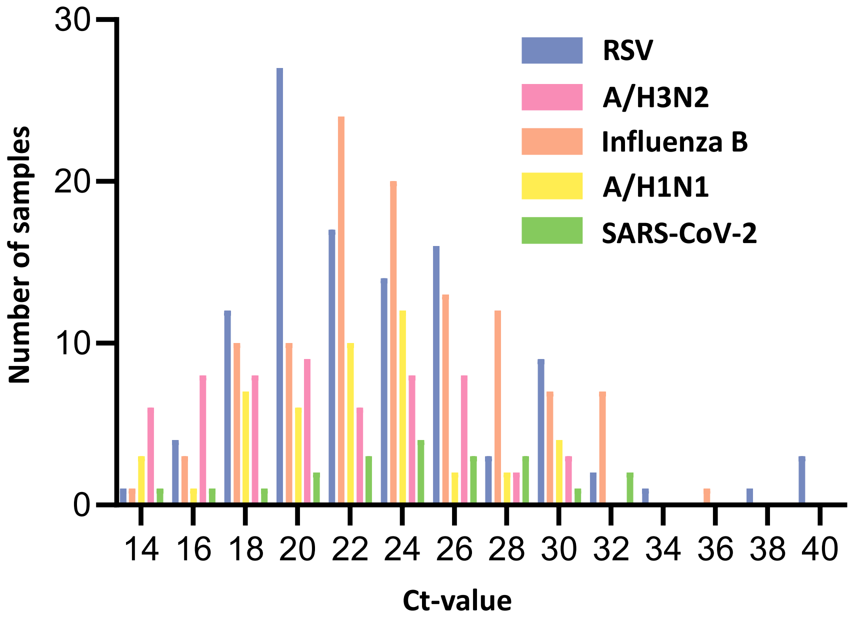

**Supplementary Figure 6. Distribution of Cycle Threshold (Ct)-values in the sample cohort.** A total of 344 clinical nasopharyngeal swabs were analyzed using the Winterplex immuno-MRM method. The Ct-values obtained through RT-qPCR ranged from Ct 12.52 to 39.88, with a median Ct of 23.24.

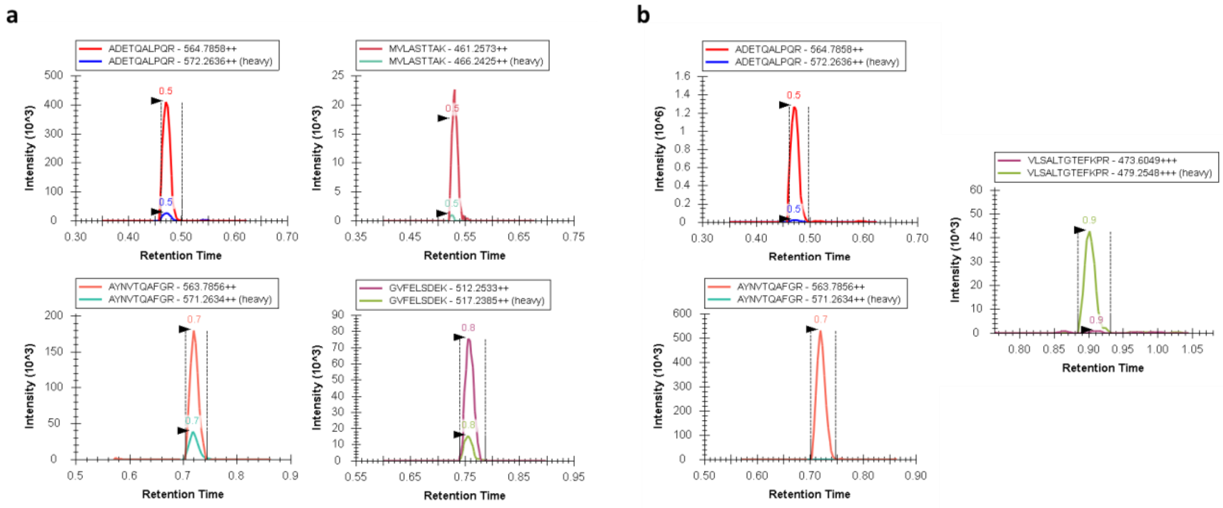

**Supplementary Figure 7.** Extracted ion chromatograms demonstrating the added resolution of the Winterplex iMRM method in detecting viral co-infections and potential misclassifications. [a] Sample 255207, classified by RT-qPCR as positive for influenza A and SARS-CoV-2, showed a clear signal for the peptide targets of both viruses, revealing a dual infection. This highlights the capability of the Winterplex iMRM to detect co-infections that might be overlooked if single-plex methodologies are used. [b] Sample 255598, classified as Influenza B by RT-qPCR, tested positive for SARS-CoV-2 with iMRM, evidenced by high signal intensities for both the ADE and AYN peptide, while no visible signal was seen for the influenza B VLS peptide, suggesting either a misclassification or an error in LIMS registration.

### Supplementary Tables

**Supplementary Table 1.** MRM parameters for the target peptides and the corresponding Polyquant QconCAT internal standard. The SIL standard peptides and their corresponding fragment ions, used for monitoring system performance, are italicized.

| Peptide | Precursor m/z | Fragment m/z | Cone Voltage (V) | Collision Energy (V) |
|---|---|---|---|---|
| **ADETQALPQR (SARS-CoV-2)** | 564.78 | 400.23 | 20 | 17 |
| | 564.78 | 584.35 | 20 | 16 |
| | 564.78 | 712.41 | 20 | 17 |
| | 572.26 | 407.21 | 20 | 17 |
| | 572.26 | 593.32 | 20 | 16 |
| | *569.79* | *410.24* | *20* | *17* |
| | *569.79* | *594.36* | *20* | *16* |
| **AYETQALPQR (SARS-CoV-2)** | 588.8 | 400.2303 | 20 | 17 |
| | 588.8 | 584.3515 | 20 | 16 |
| | 588.8 | 712.41 | 20 | 17 |
| | 596.28 | 407.2095 | 20 | 17 |
| | 596.28 | 593.3248 | 20 | 16 |
| **AYNVTQAFGR (SARS-CoV-2)** | 563.79 | 349.15 | 30 | 17 |
| | 563.79 | 679.35 | 30 | 16 |
| | 563.79 | 892.46 | 30 | 17 |
| | 571.26 | 353.14 | 30 | 17 |
| | 571.26 | 689.32 | 30 | 16 |
| | *568.79* | *689.36* | *30* | *16* |
| | *568.79* | *902.47* | *30* | *17* |
| **SMVVVRPSVASK (Influenza B)** | 420.58 | 422.26 | 30 | 13 |
| | 420.58 | 471.79 | 30 | 13 |
| | 420.58 | 521.32 | 30 | 13 |
| | 425.89 | 478.27 | 30 | 13 |
| | 425.89 | 528.3 | 30 | 13 |
| | *423.25* | *475.80* | *30* | *13* |
| | *423.25* | *525.33* | *30* | *13* |
| **VLSALTGTEFKPR (Influenza B)** | 473.6 | 524.79 | 25 | 16 |

| | | | | |
|---|---|---|---|---|
| | 473.6 | 560.31 | 25 | 16 |
| | 473.6 | 603.83 | 25 | 13 |
| | 479.25 | 531.27 | 25 | 16 |
| | 479.25 | 611.31 | 25 | 13 |
| | *476.94* | *529.80* | *25* | *16* |
| | *476.94* | *608.83* | *25* | *13* |
| **MVLASTTAK (Influenza A)** | 461.26 | 507.28 | 30 | 16 |
| | 461.26 | 578.31 | 30 | 13 |
| | 461.26 | 691.4 | 30 | 13 |
| | 466.24 | 585.29 | 30 | 13 |
| | 466.24 | 699.37 | 30 | 13 |
| | *465.26* | *586.33* | *30* | *13* |
| | *465.26* | *699.41* | *30* | *13* |
| **GVFELSDEK (Influenza A)** | 512.25 | 434.21 | 30 | 13 |
| | 512.25 | 591.3 | 30 | 16 |
| | 512.25 | 867.41 | 30 | 13 |
| | 517.24 | 438.2 | 30 | 13 |
| | 517.24 | 875.39 | 30 | 13 |
| | *516.26* | *438.22* | *30* | *13* |
| | *516.26* | *875.42* | *30* | *13* |
| **GRGVFELSDEK (Influenza A - H3N2)** | 412.88 | 478.21 | 30 | 10 |
| | 412.88 | 591.3 | 30 | 10 |
| | 412.88 | 646.33 | 30 | 14 |
| | 417.86 | 483.2 | 30 | 10 |
| | 417.86 | 655.3 | 30 | 14 |
| **VIIPTYLR (RSV)** | 487.81 | 649.37 | 20 | 13 |
| | 487.81 | 762.45 | 20 | 13 |
| | 487.81 | 552.31 | 20 | 22 |
| | 493.29 | 657.34 | 20 | 13 |
| | 493.29 | 771.42 | 20 | 13 |
| | *492.81* | *659.38* | *20* | *13* |
| | *492.81* | *772.46* | *20* | *13* |
| **DQLLSSSK (RSV)** | 439.23 | 408.21 | 25 | 16 |
| | 439.23 | 521.29 | 25 | 13 |

| | | | | |
|---|---|---|---|---|
| | 439.23 | 634.38 | 25 | 10 |
| | 444.22 | 413.19 | 25 | 16 |
| | 444.22 | 527.28 | 25 | 13 |
| | *443.24* | *416.22* | *25* | *16* |
| | *443.24* | *529.31* | *25* | *13* |

**Supplementary Table 2.** LC gradient used for Zeno SWATH analysis of the flow-through.

| Time (min) | %A | %B |
|---|---|---|
| **0** | 98.5 | 1.5 |
| **0.5** | 92.7 | 7.3 |
| **1** | 89.5 | 10.5 |
| **2** | 87 | 13 |
| **3** | 85.3 | 14.7 |
| **5** | 83.2 | 16.8 |
| **7.5** | 80.9 | 19.1 |
| **10** | 78.9 | 21.1 |
| **12.5** | 76.5 | 23.5 |
| **15** | 73.8 | 26.2 |
| **17** | 70 | 30 |
| **18** | 66.7 | 33.3 |
| **19** | 62.6 | 37.4 |
| **19.5** | 60 | 45 |
| **20** | 55 | 42 |
| **21** | 10 | 90 |
| **25** | 10 | 90 |
| **26** | 98.5 | 1.5 |
| **30** | 98.5 | 1.5 |

**Supplementary Table 3.** 85-Variable Window (VW) scheme used for the Zeno SWATH analysis of the flow-through of mRNA transfected HEK cells.

| | Start Mass (Da) | Stop Mass (Da) | CES |
|---|---|---|---|
| Exp 1 | 399.5 | 406.5 | 0 |
| Exp 2 | 405.5 | 412.5 | 0 |
| Exp 3 | 411.5 | 418.5 | 0 |
| Exp 4 | 417.5 | 424.5 | 0 |
| Exp 5 | 423.5 | 430.5 | 0 |
| Exp 6 | 429.5 | 436.5 | 0 |
| Exp 7 | 435.5 | 442.5 | 0 |
| Exp 8 | 441.5 | 448.5 | 0 |
| Exp 9 | 447.5 | 454.5 | 0 |
| Exp 10 | 453.5 | 459.5 | 0 |
| Exp 11 | 458.5 | 464.5 | 0 |
| Exp 12 | 463.5 | 469.5 | 0 |
| Exp 13 | 468.5 | 474.5 | 0 |
| Exp 14 | 473.5 | 479.5 | 0 |
| Exp 15 | 478.5 | 484.5 | 0 |

| | | | |
|---|---|---|---|
| Exp 16 | 483.5 | 489.5 | 0 |
| Exp 17 | 488.5 | 494.5 | 0 |
| Exp 18 | 493.5 | 499.5 | 0 |
| Exp 19 | 498.5 | 504.5 | 0 |
| Exp 20 | 503.5 | 509.5 | 0 |
| Exp 21 | 508.5 | 514.5 | 0 |
| Exp 22 | 513.5 | 519.5 | 0 |
| Exp 23 | 518.5 | 524.5 | 0 |
| Exp 24 | 523.5 | 529.5 | 0 |
| Exp 25 | 528.5 | 534.5 | 0 |
| Exp 26 | 533.5 | 539.5 | 0 |
| Exp 27 | 538.5 | 544.5 | 0 |
| Exp 28 | 543.5 | 549.5 | 0 |
| Exp 29 | 548.5 | 554.5 | 0 |
| Exp 30 | 553.5 | 559.5 | 0 |
| Exp 31 | 558.5 | 564.5 | 0 |
| Exp 32 | 563.5 | 569.5 | 0 |
| Exp 33 | 568.5 | 574.5 | 0 |
| Exp 34 | 573.5 | 579.5 | 0 |
| Exp 35 | 578.5 | 584.5 | 0 |
| Exp 36 | 583.5 | 589.5 | 0 |
| Exp 37 | 588.5 | 594.5 | 0 |
| Exp 38 | 593.5 | 599.5 | 0 |
| Exp 39 | 598.5 | 604.5 | 0 |
| Exp 40 | 603.5 | 609.5 | 0 |
| Exp 41 | 608.5 | 614.5 | 0 |
| Exp 42 | 613.5 | 619.5 | 0 |
| Exp 43 | 618.5 | 624.5 | 0 |
| Exp 44 | 623.5 | 629.5 | 0 |
| Exp 45 | 628.5 | 634.5 | 0 |
| Exp 46 | 633.5 | 639.5 | 0 |
| Exp 47 | 638.5 | 644.5 | 0 |
| Exp 48 | 643.5 | 649.5 | 0 |
| Exp 49 | 648.5 | 654.5 | 0 |
| Exp 50 | 653.5 | 660.5 | 0 |
| Exp 51 | 659.5 | 666.5 | 0 |
| Exp 52 | 665.5 | 672.5 | 0 |
| Exp 53 | 671.5 | 678.5 | 0 |
| Exp 54 | 677.5 | 684.5 | 0 |
| Exp 55 | 683.5 | 690.5 | 0 |

| | | | |
|---|---|---|---|
| Exp 56 | 689.5 | 696.5 | 0 |
| Exp 57 | 695.5 | 702.5 | 0 |
| Exp 58 | 701.5 | 708.5 | 0 |
| Exp 59 | 707.5 | 714.5 | 0 |
| Exp 60 | 713.5 | 720.5 | 0 |
| Exp 61 | 719.5 | 726.5 | 0 |
| Exp 62 | 725.5 | 732.5 | 0 |
| Exp 63 | 731.5 | 738.5 | 0 |
| Exp 64 | 737.5 | 744.5 | 0 |
| Exp 65 | 743.5 | 750.5 | 0 |
| Exp 66 | 749.5 | 756.5 | 0 |
| Exp 67 | 755.5 | 763.5 | 0 |
| Exp 68 | 762.5 | 770.5 | 0 |
| Exp 69 | 769.5 | 777.5 | 0 |
| Exp 70 | 776.5 | 784.5 | 0 |
| Exp 71 | 783.5 | 791.5 | 0 |
| Exp 72 | 790.5 | 798.5 | 0 |
| Exp 73 | 797.5 | 805.5 | 0 |
| Exp 74 | 804.5 | 812.5 | 0 |
| Exp 75 | 811.5 | 819.5 | 0 |
| Exp 76 | 818.5 | 826.5 | 0 |
| Exp 77 | 825.5 | 834.5 | 0 |
| Exp 78 | 833.5 | 842.5 | 0 |
| Exp 79 | 841.5 | 850.5 | 0 |
| Exp 80 | 849.5 | 858.5 | 0 |
| Exp 81 | 857.5 | 867.5 | 0 |
| Exp 82 | 866.5 | 876.5 | 0 |
| Exp 83 | 875.5 | 885.5 | 0 |
| Exp 84 | 884.5 | 894.5 | 0 |
| Exp 85 | 893.5 | 903.5 | 0 |
